# Parkinson’s disease-associated LRRK2 armadillo domain variants enhance kinase pathway activity through a Rab-binding site-dependent mechanism

**DOI:** 10.64898/2026.11.24.26368659

**Authors:** Nur Rashidova, Sara Gomes, Rose E Francis, Richard Scharff, Theresa Matheson-Grant, Pawel Lis, Raja Nirujogi, Thais Gonzalez, Amelie G Alessi, the Global Parkinson’s Genetics Program (GP2), Lara M Lange, Alexander Zimprich, Suzanne R. Pfeffer, Lydia Vela Desojo, Janet Hoenicka, Esther Sammler, Dario R Alessi

**Affiliations:** MRC Protein Phosphorylation and Ubiquitylation Unit, Faculty of Life Sciences, University of Dundee Dow Street, DD1 5EH, UK; Department of Biochemistry, Stanford University School of Medicine, Stanford, California, USA; Aligning Science Across Parkinson’s (ASAP) Collaborative Research Network, Chevy Chase, Maryland, USA; Department of Neurology, University Medical Center Hamburg-Eppendorf, Hamburg, Germany; Department of Neurology, Medical University of Vienna, Vienna, Austria; Movement Disorders Unit, Department of Neurology, Hospital Universitario Fundación Alcorcón, Madrid, Spain; Laboratory of Neurogenetics and Molecular Medicine, Institut de Recerca Sant Joan de Déu, Barcelona, Spain; Centro de Investigación Biomédica en Red de Enfermedades Raras (CIBERER), ISCIII, Madrid, Spain; Division of Neuroscience, Faculty of Health, University of Dundee, Dundee DD2 1GZ, UK

## Abstract

Pathogenic variants that increase LRRK2 kinase activity are a common genetic cause of Parkinson’s disease (PD). Most pathogenic variants occur within the catalytic ROC-COR GTPase (e.g. R1441C) and kinase (e.g. G2019S) domains, where they activate LRRK2 through distinct mechanisms. The common kinase-domain variant directly enhances the intrinsic catalytic activity of LRRK2, whereas ROC-COR variants shift the equilibrium towards its active conformation. Whether variants in the N-terminal armadillo (ARM) domain, which contains Rab GTPase-binding sites involved in LRRK2 membrane recruitment, activate LRRK2 through a different mechanism remains unclear.

Here, we characterise the LRRK2 Madrid haplotype, comprising two ARM variants (L119P/L488P) identified in a multigenerational family with PD. In cellular assays, L488P, but not L119P, increased LRRK2-mediated Rab10 phosphorylation approximately two-fold. Increased LRRK2 pathway activity was likewise observed in primary neutrophils and monocytes from variant carriers, including individuals with and without PD. Multiplexed parallel reaction monitoring (PRM) mass spectrometry in neutrophils further demonstrated coordinated increases in phosphorylation of six endogenous Rab substrates (Rab3, Rab8, Rab10, Rab12, Rab35 and Rab43), demonstrating concordant pathway activation in cellular models and primary human blood cells.

Mechanistically, L488P lies adjacent to ARM Rab-binding site 1, and its activating effect was abolished by the site-1-disrupting L403E substitution, whereas activation by the ROC-domain variant R1441C was retained. We further investigated five rare ARM-domain variants identified through GP2 and independent genetic datasets, of which two, F470S and V483G, showed increased LRRK2 pathway activity in a site-1-dependent manner, while the remaining three showed no substantial activation. Together, these findings demonstrate that L488P-mediated pathway activation is recapitulated in human variant carriers and depends on an intact ARM Rab-binding site 1, identifying a mechanism of LRRK2 regulation distinct from that of classical catalytic-core variants.

## Introduction

Parkinson’s disease (PD) is the second most common neurodegenerative disorder, and variants in *LRRK2* are one of its more frequent monogenic causes. (1, 2) Pathogenic LRRK2 variants act through a gain-of-function increase in kinase activity, (3, 4) (5) and LRRK2 kinase inhibitors are now being evaluated in clinical trials as potential disease-modifying therapies. (6–8) LRRK2 phosphorylates a subset of Rab GTPases on a conserved threonine or serine in their Switch-II motif, including Rab8A (Thr72), Rab10 (Thr73), Rab12 (Ser106), Rab35 (Thr72) and Rab43 (Thr82) (9, 10) and the levels of these phospho-Rab species provide a direct, quantitative readout of cellular LRRK2 activity that is now widely used to assess pathogenic variants and to monitor target engagement in patient samples. (5, 7, 11–13)

LRRK2 is a large multidomain protein comprising an N-terminal armadillo (ARM) domain, ankyrin (ANK) and leucine-rich repeat (LRR) domains, an atypical ROC GTPase domain, an associated COR domain (subdivided into the DEP1, DEP2, PPM-like and U-box subdomains) (5), the kinase domain and a C-terminal WD40 domain. (4) The great majority of well-characterised pathogenic variants map to the ROC-COR and kinase domains. (5) ROC-COR variants, including R1441C/G/H, N1437H and Y1699C, increase kinase activity largely by destabilising the inactive conformation of LRRK2 and thereby shifting the conformational equilibrium towards the active state (5, 14, 15), whereas the kinase-domain variants G2019S and I2020T directly enhance the intrinsic activity of the kinase. (16–18) Ser910 and Ser935 are two 14-3-3-binding sites associated with the inactive, cytosolic pool of LRRK2. Unlike kinase-domain variants, strongly activating ROC-COR variants, as well as Type I kinase inhibitors, induce dephosphorylation of Ser910 and Ser935 and dissociation of 14-3-3, reflecting changes in LRRK2 conformation and regulation. (19–22)

Beyond its catalytic core, the LRRK2 ARM domain contains at least four Rab-binding surfaces that regulate kinase recruitment and activation at membranes. (23–28) Two of these sites are well characterised: ARM Rab-binding site 1, which binds Rab29, Rab32 and Rab38 (23, 26, 29), as well as Rab8A and Rab10 (23), and site 3, which binds Rab12. (24, 25, 27) Rab-dependent recruitment of LRRK2 to damaged lysosomes and other membranes is increasingly recognised as a key step in physiological and pathological LRRK2 activation. (25, 30–33) Whether inherited variants affecting ARM Rab-binding surfaces activate LRRK2 through mechanisms distinct from classical catalytic-core variants has not been addressed.

Here, we investigate the PD-associated LRRK2 Madrid haplotype (L119P/L488P) identified in a multigenerational family with PD. (34) L488P maps immediately adjacent to ARM Rab-binding site 1 (Fig. 1A). We show that L488P, but not L119P, increased LRRK2-dependent Rab phosphorylation in peripheral blood samples from variant carriers. By integrating in vivo biomarker analysis in 14 members of this three-generation family with cell-based kinase assays and structure-guided mutagenesis, we demonstrate that L488P-mediated LRRK2 activation depends on an intact ARM Rab-binding site 1. We further identify additional rare variants near this site that increase LRRK2 pathway activity and show similar site-1 dependence. These findings extend our understanding of how ARM-domain variation influences LRRK2 activity, how the study of rare human genetic variation can uncover new biology, and highlight ARM Rab-binding site 1 as an important regulatory interface with potential therapeutic relevance in PD.

**Figure 1.**
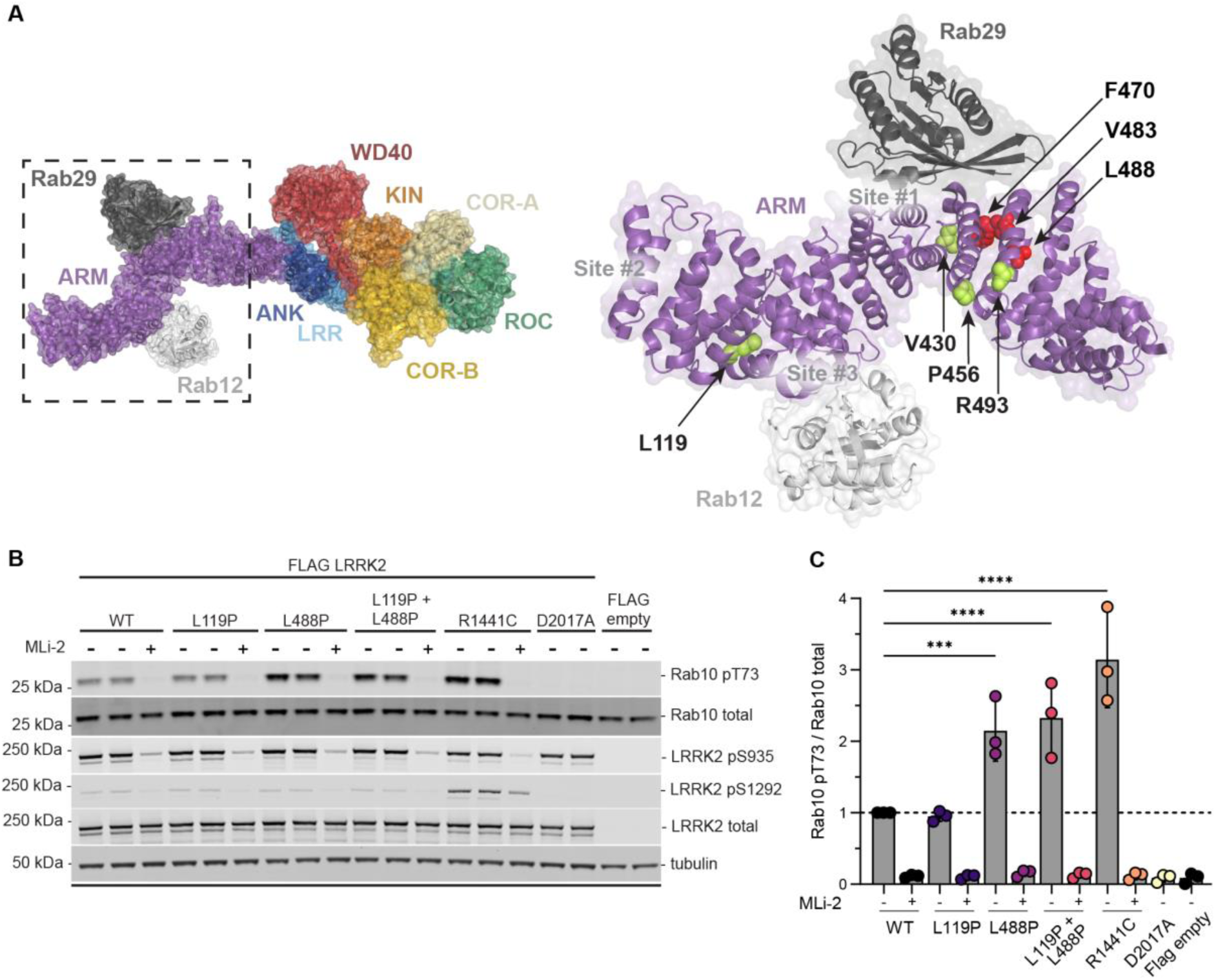
The Parkinson’s disease–associated LRRK2 L488P variant increases LRRK2-dependent phosphorylation of Rab10 at Thr73. (A) Mapping of the PD-associated LRRK2 armadillo-domain variants investigated in this study onto cryo-electron microscopy structures (PDB: 8FO2 and 8VH4), showing Rab29 and Rab12 binding at sites 1 and 3, respectively. Hyperactivating variants are shown in red and non-activating variants in green. **(B)** Functional analysis of the Madrid variants using transient overexpression of FLAG-tagged LRRK2 constructs in HEK293 LRRK2-knockout cells. Constructs comprised the Madrid haplotype (L119P/L488P), the individual L119P and L488P variants, wild-type (WT) LRRK2, the PD-associated R1441C variant (positive control), and the kinase-dead D2017A variant (negative control). An empty FLAG vector served as an additional control. Where indicated, cells were treated with 200 nM MLi-2 for 3 h before lysis. **(C)** Quantification of Rab10 Thr73 phosphorylation from three independent biological replicates, expressed as the ratio of pThr73-Rab10 to total Rab10. Values within each replicate were normalised to the mean value for WT LRRK2 in that replicate. Statistical analysis was performed using one-way ANOVA. The dashed horizontal line indicates the normalised WT level (1.0). The immunoblot shown is representative of three independent biological replicates. Source data are available on Zenodo (DOI: 10.5281/zenodo.23360970).

## Results

### The Parkinson’s-associated LRRK2 L488P substitution hyperactivates the kinase

The L119P and L488P substitutions map to the N-terminal armadillo (ARM) domain of LRRK2 (Fig. 1A). L119P lies distal to the two well-characterised Rab-binding sites of the ARM domain: site 1, which binds Rab29, Rab32 and Rab8A/Rab10 (23, 26), and site 3, which binds Rab12. (24, 25, 27) In contrast, L488P lies immediately adjacent to Rab-binding site 1, within a long α-helix that flanks but does not overlap the binding surface (Fig. 1A). Because proline substitutions are predicted to disrupt α-helical structure, L488P could distort this helix and thereby perturb the conformation of the neighbouring Rab-binding site 1.

To test whether these substitutions affect kinase activity, we transiently expressed the LRRK2 L119P/L488P variant in LRRK2-knockout HEK293 cells and compared its activity with wild-type LRRK2. LRRK2 kinase pathway activity was assessed by immunoblotting for phosphorylation of Rab10 at Thr73 using phospho-specific antibodies, a well-established assay used to evaluate PD-associated LRRK2 variants. (5, 15) Relative to wild-type LRRK2, the L119P/L488P variant increased Rab10 phosphorylation more than two-fold, approaching, though remaining slightly below, the strongly activating LRRK2 R1441C variant, which increased activity approximately three-fold in the same experiments (Fig. 1B,C). Treatment with the LRRK2 inhibitor MLi-2 reduced Rab10 phosphorylation to undetectable levels in cells expressing wild-type LRRK2, L119P, L488P or the L119P/L488P double variant, confirming that the observed phosphorylation was LRRK2 kinase-dependent. As expected, the kinase-dead LRRK2 D2017A variant showed no detectable Rab10 phosphorylation (Fig. 1B,C).

To identify which substitution was responsible, we tested the two variants individually. L119P displayed activity comparable to wild-type LRRK2, whereas L488P increased Rab10 phosphorylation more than two-fold, an increase indistinguishable from that of the L119P/L488P double variant (Fig. 1B,C). The elevated activity of the double variant is therefore attributable to L488P alone. LRRK2 expression levels were comparable across wild-type and the L119P, L488P and double variants. LRRK2 autophosphorylation at Ser1292 was most prominent in the R1441C positive control, with no obvious increase in the L488P-containing variants compared with wild-type LRRK2. The pSer1292 signal was reduced following treatment with MLi-2 (Fig. 1B). Phosphorylation of Ser935, a 14-3-3-binding site associated with the inactive, cytosolic conformation of LRRK2 (21, 22), was unchanged by any of the variants (Fig. 1B,C), suggesting that L488P-driven activation does not involve the pronounced changes in LRRK2 regulation associated with strongly activating ROC-COR variants. Together with the proximity of L488P to Rab-binding site 1, these findings suggest that activation results from a local effect on the ARM domain rather than a global shift in LRRK2 conformation.

### L119P/L488P carriers show elevated LRRK2-dependent Rab phosphorylation in peripheral blood monocytes and neutrophils

Having established that L488P activates LRRK2 in a cellular overexpression system, we next asked whether this translated into increased pathway activity *in vivo* by analysing peripheral blood neutrophils and monocytes from 14 members of a three-generation family, including both manifesting and non-manifesting L119P/L488P variant carriers (Fig. 2). These cell populations provide a robust system for assessing human LRRK2 pathway activity, as they can be readily isolated from fresh blood, express abundant endogenous LRRK2 and have been used extensively to evaluate the effects of LRRK2 variants in humans. (5, 11, 35–39) Genetic testing identified nine carriers of the L119P/L488P haplotype, including two individuals with PD, and five non-carrier family members.

**Figure 2.**
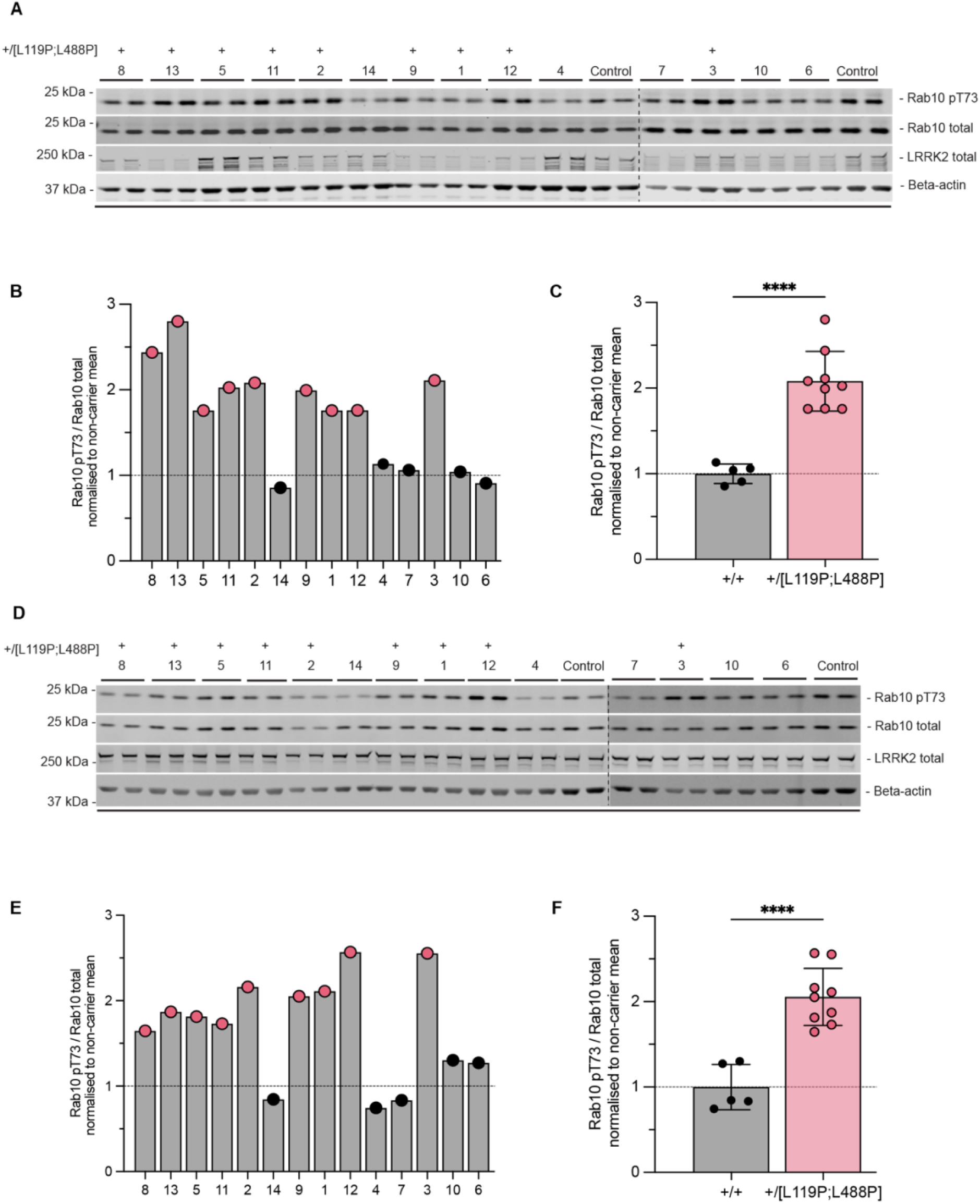
Increased LRRK2-dependent Rab10 phosphorylation in neutrophils and monocytes from LRRK2 L119P;L488P carriers. **(A)** Immunoblot analysis of neutrophil lysates from 14 members of the Madrid family, comprising nine LRRK2 L119P;L488P haplotype carriers and five non-carrier family members. Lysates (15 µg total protein per participant) were loaded in duplicate and analysed using the indicated antibodies. **(B)** Quantification of Rab10 Thr73 phosphorylation in neutrophils, expressed as the ratio of pThr73-Rab10 to total Rab10 and normalised to the mean of the non-carrier family members. Each bar represents the mean of duplicate loadings from one participant. **(C)** Grouped analysis of neutrophil Rab10 Thr73 phosphorylation according to LRRK2 genotype. **(D)** Immunoblot analysis of monocyte lysates from the same 14 family members. Lysates (15 µg total protein per participant) were loaded in duplicate and analysed using the indicated antibodies. **(E)** Quantification of Rab10 Thr73 phosphorylation in monocytes, expressed as the ratio of pThr73-Rab10 to total Rab10 and normalised to the mean of the non-carrier family members. Each bar represents the mean of duplicate loadings from one participant. **(F)** Grouped analysis of monocyte Rab10 Thr73 phosphorylation according to LRRK2 genotype. L119P;L488P haplotype carriers are shown in pink and non-carriers in black. Each bar in **(B)** and **(E)** and each point in **(C)** and **(F)**, represents one participant and is the mean of duplicate technical loadings. Data are shown as mean ± SD. Groups were compared using unpaired, two-tailed Welch’s t-tests. ****P ≤ 0.0001. Dashed horizontal lines indicate the normalised non-carrier mean (1.0). Participant numbers were randomly reassigned for figure presentation and do not correspond to the original sample identifiers. Identical numbers indicate paired neutrophil and monocyte samples from the same participant. (Source data are available on Zenodo; DOI: 10.5281/zenodo.23360970).

LRRK2-dependent Rab phosphorylation in neutrophils (Fig. 2A-C) and monocytes (Fig. 2D-F) was assessed by phospho-Rab10 immunoblotting. This revealed an approximately two-fold increase in Rab10 phosphorylation in both cell populations from L119P/L488P variant carriers compared with non-carrier family members, with all nine carriers displaying higher phospho-Rab10 levels than each of the five non-carriers (Fig. 2C,F).

To independently quantify LRRK2 pathway activity across multiple endogenous Rab substrates, we also analysed neutrophil samples using multiplexed targeted parallel reaction monitoring (PRM) mass spectrometry (Fig. 3). To expand the range of detectable LRRK2 substrates, we first evaluated a newly identified phospho-Rab12 (Ser106) immunoprecipitating antibody (Abcam AB325632; clone MJF-D29764-507; BSA- and azide-free), which efficiently captured the LRRK2-phosphorylated Rab12 tryptic peptide (Supplementary Fig. 1A). We subsequently incorporated this antibody into our previously optimized, automated multiplexed PRM workflow, enabling simultaneous quantification of LRRK2-dependent phosphorylation of Rab1, Rab3, Rab8, Rab10, Rab12, Rab35 and Rab43 in clinical samples. The workflow builds on a previously published method (40), with the updated protocol available on protocols.io (https://dx.doi.org/10.17504/protocols.io.5jyl839y8v2w/v1(41); manuscript in preparation).

**Figure 3.**
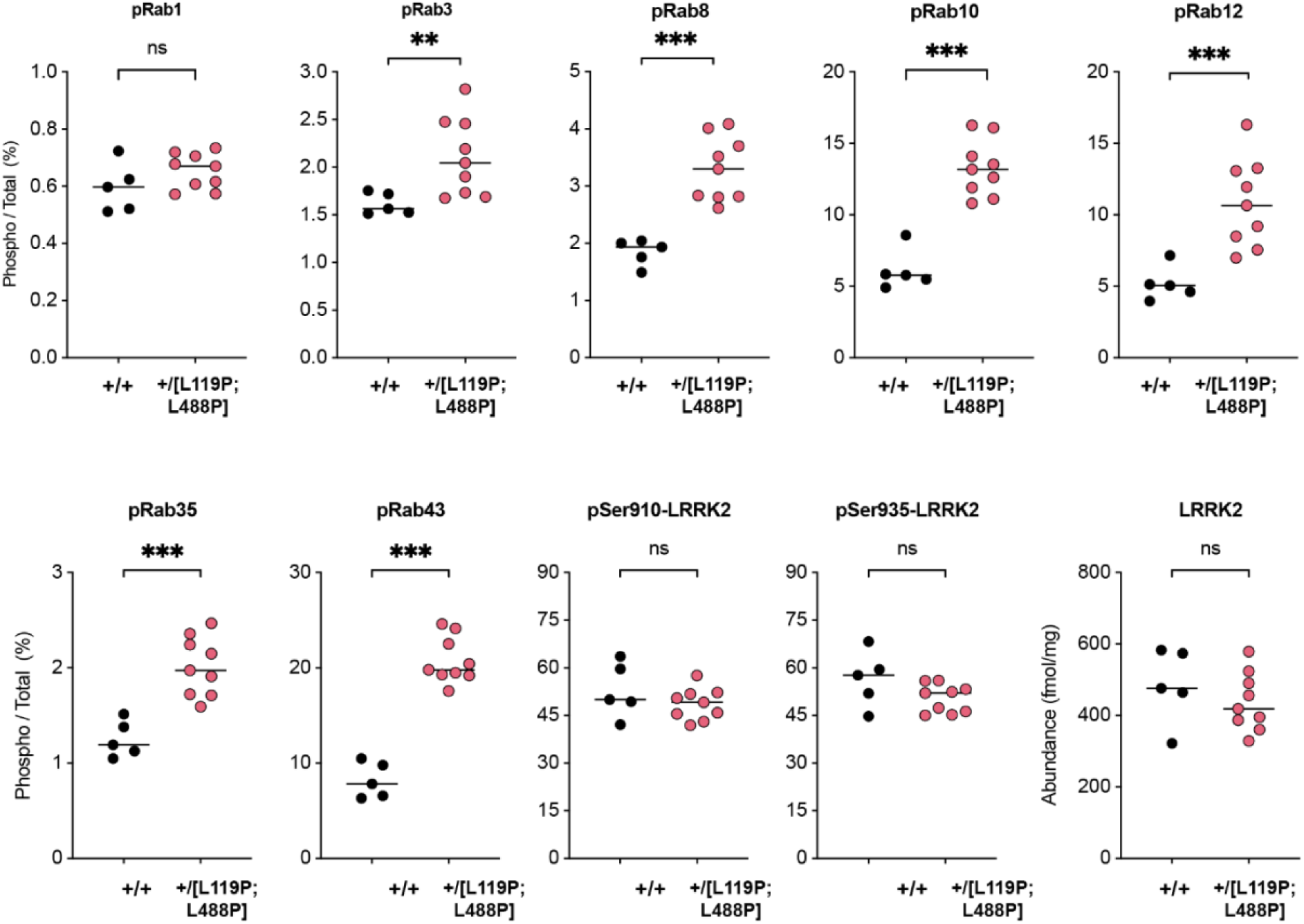
Multiplexed PRM mass spectrometry reveals increased LRRK2-dependent Rab phosphorylation in peripheral blood neutrophils from LRRK2 L119P; L488P carriers. Neutrophils isolated from fresh peripheral blood from members of the Madrid family carrying the LRRK2 L119P;L488P haplotype (n = 9) and non-carrier family members (n = 5) were analysed using a multiplexed parallel reaction monitoring (PRM) mass spectrometry assay. Phosphorylated and corresponding total Rab peptides were quantified for Rab1, Rab3, Rab8, Rab10, Rab12, Rab35, and Rab43. Rab phosphorylation is expressed as the abundance of the phosphorylated peptide relative to the corresponding total Rab peptide (%). Total LRRK2 abundance is expressed as fmol/mg protein and LRRK2 phosphorylation at Ser910 and Ser935 was quantified and expressed as phosphorylation relative to total LRRK2 (%). Each point represents an individual participant, and horizontal lines indicate the mean. Groups were compared using two-sided unpaired Welch’s t-tests on log10-transformed data, with Benjamini-Hochberg correction for multiple comparisons (FDR < 0.05). Significance annotations represent FDR-adjusted q values: q < 0.05 (*), q <0.01 (**), q < 0.001 (***), ns, not significant. Exact unadjusted P values and FDR-adjusted q values are provided in Supplementary Table 1. Source data are available on Zenodo (DOI: 10.5281/zenodo.23360970). The mass spectrometry proteomics data have been deposited with the ProteomeXchange Consortium via the PRIDE repository under dataset identifier PXD085751.

Consistent with the immunoblotting findings, multiplexed PRM analysis demonstrated coordinated increases in phosphorylation across multiple endogenous LRRK2 substrates. Relative to non-carriers, L119P/L488P variant carriers exhibited approximately 1.7-fold higher phosphorylation of Rab8 and Rab35, two-fold higher phosphorylation of Rab10 and Rab12, and 2.5-fold higher phosphorylation of Rab43 (Fig. 3). A more modest, approximately 1.3-fold increase was observed for phospho-Rab3, with all six increases remaining significant after correction for multiple testing (Benjamini–Hochberg FDR-adjusted q < 0.01; Supplementary Table 1), whereas no significant difference was observed for phospho-Rab1. Neither total LRRK2 abundance nor phosphorylation at Ser910 and Ser935 differed significantly between carriers and non-carriers. Together, these findings demonstrate that the LRRK2 pathway activation conferred by L488P in the cellular overexpression system is recapitulated in primary neutrophils and monocytes from both manifesting and non-manifesting variant carriers.

### Disruption of ARM-Rab binding site 1 suppresses LRRK2 activation by L488P

Given the proximity of L488P to ARM Rab-binding site 1, we investigated whether its activating effect depends on an intact Rab-binding interface. To test this, we introduced the L403E substitution, which abolishes binding of multiple Rab proteins to site 1(26), into the L488P variant and measured Rab10 phosphorylation in LRRK2-knockout HEK293 cells. As before, L488P increased Rab10 phosphorylation more than two-fold relative to wild-type LRRK2 (Fig. 4A,B). L403E reduced Rab10 phosphorylation by approximately 30–40% of wild-type levels, and the L403E/L488P double variant was indistinguishable from L403E alone (Fig. 4A,B).

**Figure 4.**
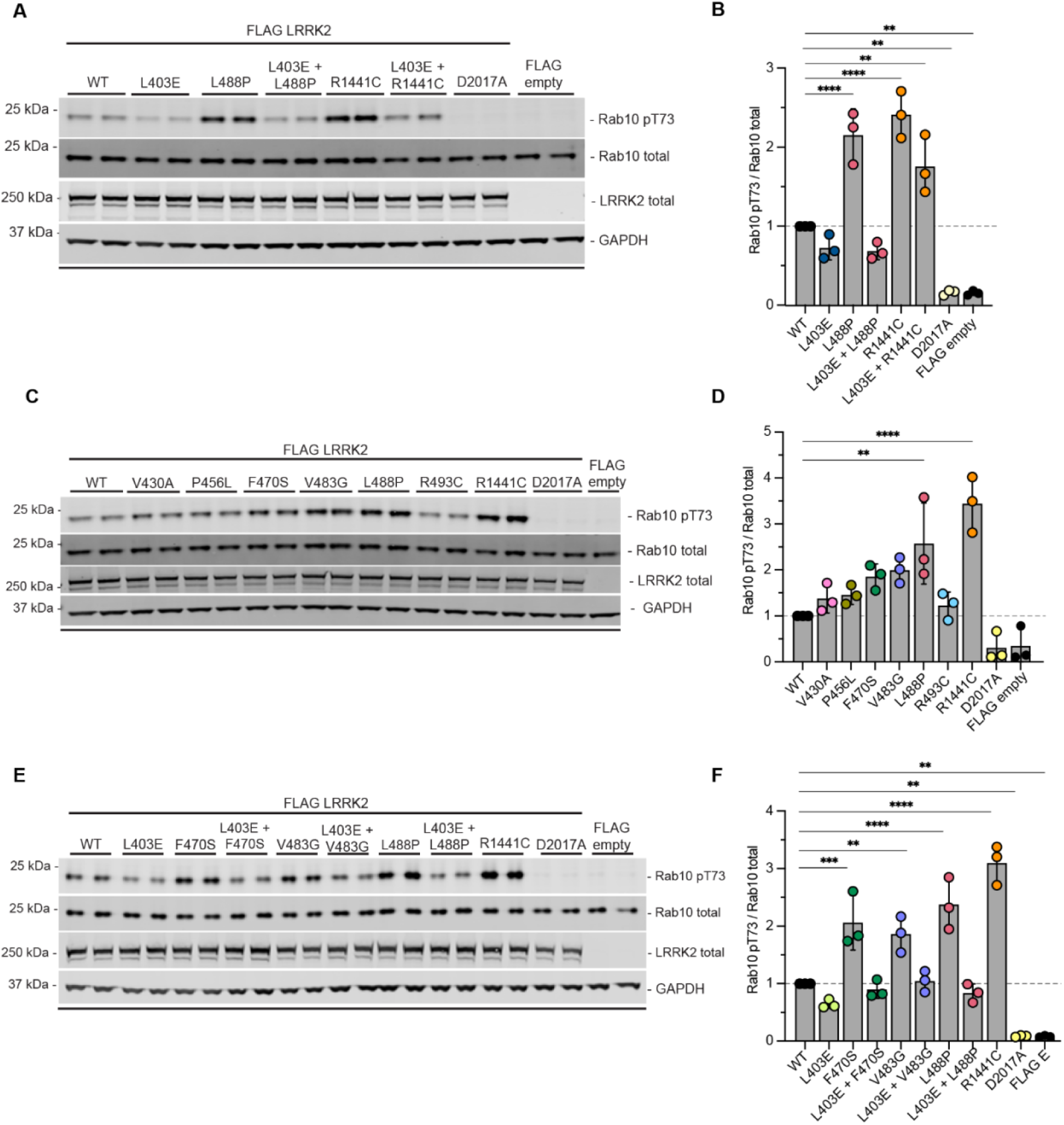
LRRK2 ARM-domain variants enhance kinase activity through a Rab-binding site 1-dependent mechanism. [A, B] Functional analysis and quantification of the L488P variant and the Rab-binding site 1-disrupting L403E substitution following transient overexpression of FLAG-tagged LRRK2 constructs in HEK293 LRRK2-knockout cells. L403E suppresses L488P-mediated LRRK2 activation. [C, D] Functional analysis and quantification of L488P and five neighbouring ARM-domain variants (V430A, P456L, F470S, V483G and R493C). [E, F] Functional analysis and quantification of the F470S, V483G and L488P variants in the presence or absence of L403E, demonstrating suppression of variant-mediated LRRK2 activation by disruption of Rab-binding site 1. R1441C and D2017A served as positive and kinase-dead controls, respectively, with empty FLAG vector as an additional negative control. Rab10 Thr73 phosphorylation was quantified as pThr73-Rab10/total Rab10 and normalised within each experiment to WT LRRK2. Data are mean ± SD from three independent biological replicates; immunoblots are representative of these experiments (Zenodo repository, DOI: 10.5281/zenodo.23360970). Statistical analysis used one-way ANOVA with Dunnett’s multiple-comparisons test versus WT. P < 0.05 (*), < 0.01 (**), < 0.001 (***) and < 0.0001 (****); non-significant comparisons are not shown. Dashed lines indicate the normalised WT level (1.0).

Disruption of site 1 therefore suppressed L488P-mediated activation completely. Because L403E also lowers basal pathway activity, we considered whether this suppression might simply reflect a general reduction in Rab10 phosphorylation rather than a specific requirement for site 1. The R1441C control argues against this interpretation. Introducing L403E into R1441C reduced basal activity to a comparable extent, yet R1441C retained clearly elevated Rab10 phosphorylation relative to L403E alone (Fig. 4A,B). A lowered basal output is therefore compatible with an activating variant still exerting its effect, and the complete loss of L488P-mediated activation on the same background cannot be attributed to the reduced starting point. This is consistent with R1441C acting primarily by shifting the equilibrium between active and inactive conformational states (18) rather than through site-1 Rab binding. Together, these findings demonstrate that L488P-mediated LRRK2 activation requires an intact ARM Rab-binding site 1.

### Identification of LRRK2 variants adjacent to L488P that enhance kinase activity

To investigate whether other variants located near ARM Rab-binding site 1 similarly increase LRRK2 pathway activity, we interrogated the GP2 (42) database and scientific literature for LRRK2 variants located near L488P. This identified five candidates for functional analysis, V430A, P456L, F470S, V483G and R493C (Fig. 4C,D). We assessed the effect of each variant on pathway activity in the LRRK2-knockout HEK293 expression system. Two of the five variants exceeded the previously established functional threshold of 1.5-fold relative to wild-type LRRK2 (5, 15): F470S increased kinase activity approximately 1.8-fold and V483G approximately 2-fold (Fig. 4C,D). However, neither increase reached statistical significance following Dunnett’s multiple-comparisons correction (adjusted P = 0.136 and P = 0.060, respectively). V483G was identified in an individual with PD (age at onset in their forties) recruited through a previously described genetic diagnostic cohort. (43) The variant is exceptionally rare in gnomAD v4 (allele count = 1) (44), and this individual has not previously been reported in detail. The F470S variant was identified in an individual with PD (age at onset in their seventies) through GP2. (42) The remaining three variants, V430A, P456L and R493C, showed no substantial increase in LRRK2 activity relative to wild-type (Fig. 4C,D). As for L488P, introduction of the site-1-disrupting L403E substitution reduced the increased Rab10 phosphorylation associated with both F470S and V483G (Fig. 4E,F), indicating that the effects of both variants depend on an intact ARM-Rab binding site 1. We also combined the V483G and L488P variants, but this did not further increase LRRK2-mediated Rab10 phosphorylation, suggesting that they act through a similar mechanism (Supplementary Fig 2). Together, these findings identify additional rare LRRK2 variants near the ARM Rab-binding site 1 with evidence of increased LRRK2 pathway activity and site 1 dependence, while demonstrating that proximity to this binding site alone is insufficient to enhance LRRK2 activity.

## Discussion

Here, we identify a distinct mechanism by which a PD-associated LRRK2 variant can increase kinase pathway activity. The ARM-domain substitution L488P, but not the co-inherited L119P variant, increased LRRK2-dependent Rab phosphorylation in HEK293 cells (Fig. 1), consistent with the elevated pathway activity observed in peripheral blood neutrophils (Figs. 2 and 3) and monocytes (Fig. 2) from L119P/L488P carriers. Several findings indicate that L488P acts through a mechanism involving ARM Rab-binding site 1. L488P lies immediately adjacent to this site and requires an intact site 1 for its activating effect (Fig. 4): disruption of site 1 by L403E completely suppressed activation by L488P. In contrast, although L403E also reduced Rab10 phosphorylation in the R1441C background, R1441C retained elevated activity relative to L403E alone. These findings support distinct mechanisms of LRRK2 activation, with L488P acting through a site-1-dependent mechanism and R1441C retaining an activating effect despite disruption of this interface. Together, these findings support a model in which L488P increases LRRK2 activity through a mechanism dependent on ARM Rab-binding site 1, revealing a mechanism of pathogenic activation distinct from the altered intrinsic kinase activity or conformational equilibrium associated with classical catalytic-core variants.

One possible explanation is that L488P alters the interaction of LRRK2 with Rab proteins engaging ARM site 1, potentially increasing its recruitment to Rab-positive membranes. Enhanced recruitment would in turn be expected to elevate basal phosphorylation of LRRK2 Rab substrates. Although this mechanism remains to be established directly, the increased phosphorylation of Rab3, Rab8, Rab10, Rab12, Rab35 and Rab43 observed in neutrophils from variant carriers is consistent with this model and provides evidence that the enhanced LRRK2 pathway activity observed in the HEK293 overexpression system is recapitulated at endogenous LRRK2 levels in primary human cells.

Beyond L488P, additional variants identified in single individuals with PD in proximity to site 1 (F470S and V483G) also exceeded the predefined activation threshold, although neither increase reached statistical significance. Mechanistically, we found that both variants required an intact site 1 for their activating effects. Together, these findings provide cell-based functional evidence that variants clustered around ARM Rab-binding site 1 can activate LRRK2 through a shared site-1-dependent mechanism. Importantly, these variants were selected for functional investigation solely based on their structural proximity to L488P, rather than genetic evidence supporting an association with PD. Neither F470S nor V483G is currently established as a disease-associated variant, and their identification in only a single individual with PD each does not establish pathogenicity or a causal relationship with disease. Thus, while these findings provide insights into the functional relevance of this region, further genetic, clinical, and functional studies are required to determine their potential contribution to PD pathogenesis.

These observations suggest that variants affecting the ARM Rab-binding interface can increase LRRK2 pathway activity through a mechanism distinct from that of classical catalytic-core variants. Together, our findings provide functional evidence supporting the importance of ARM Rab-binding site 1 in the regulation of LRRK2 activity and its potential relevance to PD.

Recent work has identified approximately 10 additional activating variants within the LRRK2 ARM domain. (5) It will be important to determine whether these variants similarly act by enhancing Rab binding or by altering interactions with other regulators that promote LRRK2 membrane recruitment. Understanding the precise biochemical effects of these variants is particularly important given the incomplete penetrance of pathogenic LRRK2 variants, with many carriers remaining asymptomatic, as illustrated by the family studied here. Independent genetic support for the importance of ARM Rab-binding site 1 comes from the recent identification of the PD-associated Rab32 S71R variant. Rab32 engages ARM site 1, supporting a role for this interface in the regulation of LRRK2 activity. (45, 46) Rab29, which also interacts with ARM site 1 and recruits LRRK2 to the Golgi, has also been genetically linked to PD. (47, 48) Together, these observations emphasise the ARM Rab-binding interface as a functionally important region in which variants with increased LRRK2 pathway activity can arise. Future work should map activating ARM-domain variants onto the Rab-binding surfaces recently characterised by cryo-electron microscopy (26–28), as well as the GABARAP-binding site that recruits LRRK2 in macrophages following activation of the STING-CASM pathway at lysosomes. (28, 30, 49) Variants located within or close to these interfaces could then be prioritised for functional analysis using the assays described here. This approach may reveal how individual variants stimulate LRRK2 activity, clarify their contribution to PD, and define the physiological importance of distinct ARM-domain Rab-binding sites. Determining how these variants influence kinase activation and downstream lysosomal pathways may also help explain why some carriers develop PD while others remain asymptomatic, providing insights into incomplete penetrance and potential mechanisms of resilience.

Our findings also have translational implications. The increased LRRK2 pathway activity associated with L488P, both alone and in combination with L119P, was abolished by the selective LRRK2 kinase inhibitor MLi-2 (Fig. 1B,C). These findings support the potential therapeutic relevance of LRRK2 kinase inhibition for carriers of the Madrid haplotype. In principle, the ARM–Rab interface itself might also be druggable, a hypothesis being pursued through the Michael J. Fox Foundation funded LRRK2 Investigative Therapeutics Exchange (LITE) initiative. (8) Moreover, the L488P variant may provide a useful cellular model for evaluating compounds targeting the ARM Rab-binding interface, as its activating effect depends on an intact site 1.

Beyond these therapeutic considerations, our findings also support the potential of phospho-Rab measurements as biomarkers of LRRK2 pathway activity. The detection of elevated phospho-Rab species in carrier neutrophils using a multiplexed, semi-automated mass spectrometry workflow, extended here with a validated phospho-Rab12 (Ser106) immunoprecipitating antibody (Supplementary Fig. 1), supports the use of such assays to identify pathway-activated individuals and for pharmacodynamic monitoring in clinical trials (Gomes et al in preparation). (41)

Several limitations should be noted. The magnitude of activation conferred by the ARM-domain variants was moderate (roughly two-fold) and lower than the stronger activating ROC-COR variants such as R1441C, and much of the mechanistic analysis relied on transient overexpression. Although our mutagenesis experiments demonstrate that L488P-mediated activation depends on an intact ARM Rab-binding site 1, we have not directly established whether L488P alters Rab-binding affinity or LRRK2 membrane recruitment. Further validation in endogenous knock-in models, together with biochemical and structural studies of ARM–Rab interactions, will be important to substantiate the proposed mechanism. Our data also do not establish which physiological Rab protein or proteins contribute to the increased activity of the L488P variant.

The clinical observations are also limited by the small cohort, comprising a single family and individual carriers of F470S and V483G. Consequently, penetrance, age at onset and the downstream cellular consequences (for example effects on lysosomal function and ciliogenesis) remain to be defined.

An additional consideration is the potential contribution of GBA1 variation to disease expression within the family, given its established role as a genetic risk factor and modifier of PD. The GBA1 variant was not consistently co-inherited with the LRRK2 haplotype. Furthermore, our previous experiments comparing wild-type and GBA1-knockout A549 cells did not identify an effect of GBA1 loss on LRRK2-dependent Rab10 phosphorylation. (50) Although context-dependent effects cannot be excluded, these findings suggest that GBA1 variation is unlikely to account for the increased LRRK2 pathway activity observed in the present study.

Further studies, including the identification of additional variants that act through a site-1-dependent mechanism, will help establish the contribution of ARM-domain Rab binding to LRRK2 activation in PD and inform future therapeutic strategies.

## Materials & Methods

### Identification and selection of LRRK2 ARM domain variants

Whole-genome and clinical exome sequencing data from the Global Parkinson’s Genetics Program (GP2), Release 10 (https://doi.org/10.5281/zenodo.15748014), were screened for rare missense variants in the LRRK2 armadillo (ARM) domain. GP2 genomic data were processed using the established quality control (QC) pipeline, as previously described. (51) In addition, we considered variants identified in independent PD cohorts, including V483G, which was identified in an individual with PD from an Austrian genetic diagnostic cohort previously described by Zimprich and colleagues. (43) Candidate variants were prioritised for functional characterisation based on their spatial proximity to LRRK2 residue L488 and ARM Rab-binding site 1 and identification in individuals with PD. Available clinical and phenotypic information was reviewed to identify variants observed in individuals with PD.

### Structural Mapping of LRRK2 Variants

Structural mapping was performed based on existing high-resolution Cryo-EM structures PDB:8FO2 (Rab29, Site #1) and PDB:8VH4 (Rab12, Site #3). Structural visualization software used is PyMOL 3.0.

### Reagents

The MLi-2 LRRK2 inhibitor was synthesized by MRC PPU Reagents and Services at the University of Dundee (https://mrcppureagents.dundee.ac.uk).

### Plasmids

All plasmids used in this study were obtained from the MRC PPU Reagents and Services and these are listed in Table 1. All constructs were sequenced using Plasmidsaurus (https://plasmidsaurus.com).

**Table 1.**
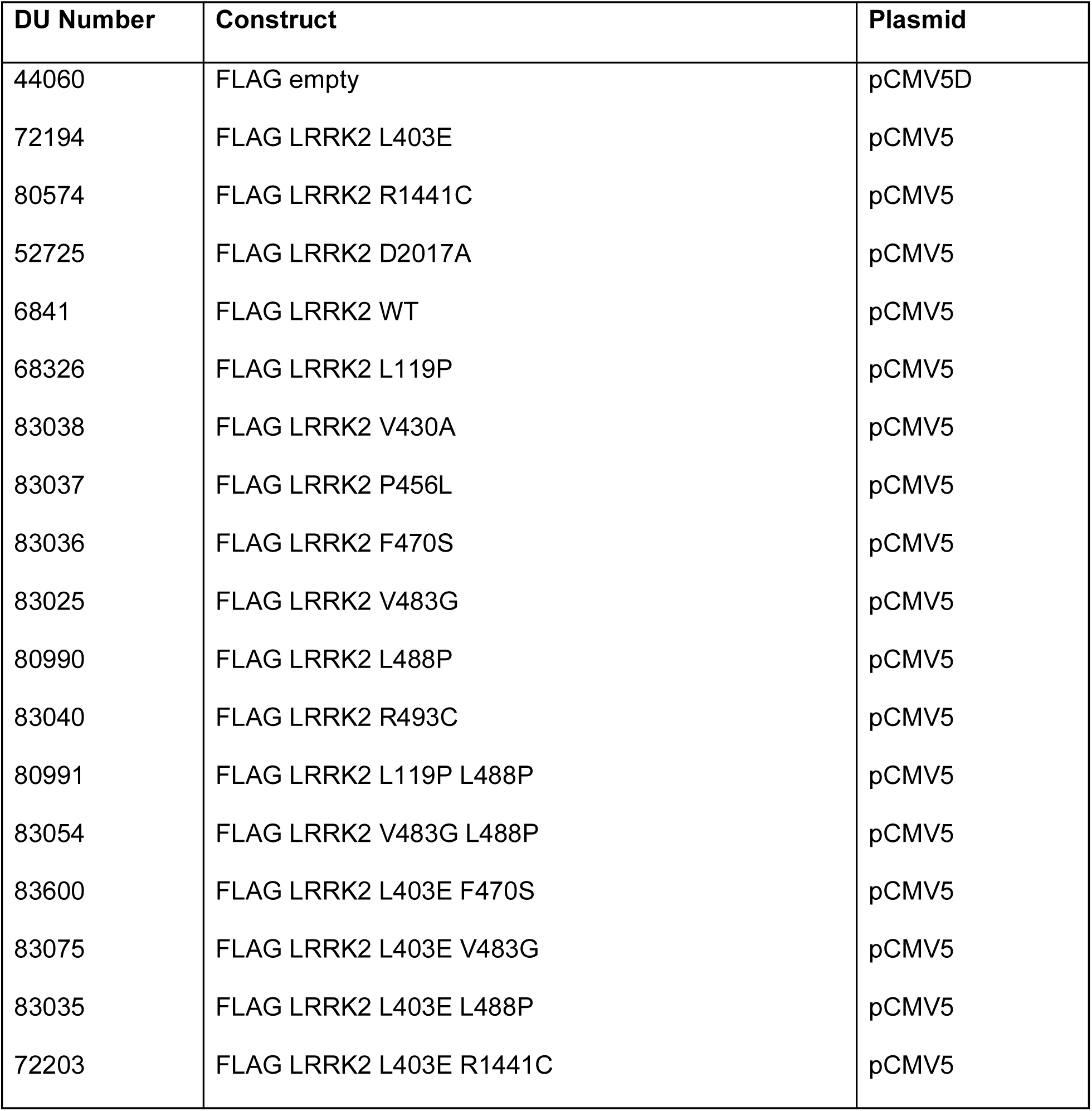
List of plasmids used in this study.

| **DU Number** | **Construct** | **Plasmid** |
|---|---|---|
| 44060 | FLAG empty | pCMV5D |
| 72194 | FLAG LRRK2 L403E | pCMV5 |
| 80574 | FLAG LRRK2 R1441C | pCMV5 |
| 52725 | FLAG LRRK2 D2017A | pCMV5 |
| 6841 | FLAG LRRK2 WT | pCMV5 |
| 68326 | FLAG LRRK2 L119P | pCMV5 |
| 83038 | FLAG LRRK2 V430A | pCMV5 |
| 83037 | FLAG LRRK2 P456L | pCMV5 |
| 83036 | FLAG LRRK2 F470S | pCMV5 |
| 83025 | FLAG LRRK2 V483G | pCMV5 |
| 80990 | FLAG LRRK2 L488P | pCMV5 |
| 83040 | FLAG LRRK2 R493C | pCMV5 |
| 80991 | FLAG LRRK2 L119P L488P | pCMV5 |
| 83054 | FLAG LRRK2 V483G L488P | pCMV5 |
| 83600 | FLAG LRRK2 L403E F470S | pCMV5 |
| 83075 | FLAG LRRK2 L403E V483G | pCMV5 |
| 83035 | FLAG LRRK2 L403E L488P | pCMV5 |
| 72203 | FLAG LRRK2 L403E R1441C | pCMV5 |

### Cell Culture

HEK293 cells (ATCC Cat no. CRL-1573, RRID:CVCL_0045) were used to create a LRRK2 KO cell line. CRISPR/Cas9 guide plasmids targeting exon 10 of the human *LRRK2* gene were designed by Thomas Macartney in MRC PPU Reagents and Services (https://mrcppureagents.dundee.ac.uk). Cell line available on the MRC PPU Intranet Cell Line Database -Cell Line ID: LRRK2 KO HEK 293 [1], Clone Number: cl D5. The sense guide plasmid was pBABED-U6-LRRK2 exon 10 KO sense (DU52443), and the antisense guide plasmid was LRRK2 exon 10 KO antisense guide + Cas9 D10A (DU52445). LRRK2 KO cells were confirmed using immunoblotting and were also confirmed using MiSeq by the MRC PPU Reagents and Services. Cells were cultured in DMEM (Dulbecco’s Modified Eagle Medium, Gibco™) containing 10% (v/v) fetal bovine serum, 2 mM l-glutamine, 100 U/ml penicillin and 100 μg/ml streptomycin at 37°C in a humidified incubator maintaining 5% (v/v) CO_2_. Cells were regularly tested for mycoplasma contamination.

### Cell Transfection and Lysis

For LRRK2 kinase activity assays, HEK293 LRRK2 KO cells were seeded in six-well plates. The following day, when cells reached 60–80% confluency, plasmids were transiently transfected using polyethyleneimine (PEI). For each well, 2 μg DNA and 6 μg PEI were diluted in 200 μL Opti-MEM™ Reduced Serum Medium (Gibco™) and added dropwise to the cells. After 24 h of transfection, cells were lysed in an ice-cold lysis buffer. The lysis buffer contains 50 mM Tris-HCl (pH 7.4), 10 mM sodium glycerophosphate, 10 mM sodium pyrophosphate, 150 mM NaCl, and 10% glycerol, and was supplemented immediately before use with 1% Triton, Complete EDTA-free protease inhibitor cocktail, and phosphatase inhibitors. Lysates stored at -80 °C. Detailed protocols are available here: https://dx.doi.org/10.17504/protocols.io.byl42ed72vo5/v1.

### Western Immunoblotting

A protocols.io description of our quantitative immunoblotting protocol has previously been described (Francesca Tonelli, Dario Alessi 2023. Quantitative Immunoblotting Analysis of LRRK2 Signalling Pathway) and is available here: https://dx.doi.org/10.17504/protocols.io.ewov14znkvr2/v2). Briefly, lysates were thawed and protein concentrations quantified using a Bradford assay (Protein Assay Dye Reagent, Bio-Rad). Lysate were adjusted to a final concentration of (2 µg/ µl) using lysis-buffer and 4x LDS loading buffer 5% (v/v) Beta-mercaptoethanol and heated at 95°C for 5 min. Samples were loaded onto NuPAGE 4–12% Bis–Tris Midi Gels (Thermo Fisher Scientific, Cat no. WG1402BOX or Cat no. WG1403BOX) and electrophoresed at 120 V for approximately 2 h with NuPAGE MOPS SDS running buffer (Thermo Fisher Scientific, Cat no. NP0001-02). At the end of electrophoresis, proteins were electrophoretically transferred onto a nitrocellulose membrane (GE Healthcare, Amersham Protran Supported 0.45 µm NC) at 90 V for 90 min on ice in transfer buffer (48 mM Tris base and 39 mM glycine supplemented with 20% (v/v) methanol). The membranes were blocked with 5% (w/v) skim milk powder dissolved in 1x TBS-T (20 mM Tris base, 150 mM sodium chloride (NaCl), 0.1% (v/v) Tween 20) at room temperature for 1 h.

Membranes were washed three times with TBS-T and were incubated in primary antibodies (diluted in 5% BSA, 1x TBS-T) overnight at 4°C. Membranes were washed three times for 15-minutes each with 1x TBST, then incubated with secondaries (diluted in 5% milk, 1x TBST) for 1 h in foil at room temperature. Thereafter, membranes were washed with 1xTBS-T three times for 15-minutes, and protein bands were acquired via near-infrared fluorescent detection using the Odyssey CLx imaging system and quantified using Image Studio Lite (Version 5.2.5, RRID:SCR_013715).

### Antibodies for Immunoblotting

**Table 2.** List of Primary Antibodies used in this study.

| Antibody target | Company | Catalogue number (RRID) | Host species | Dilution |
|---|---|---|---|---|
| LRRK2 (C-terminus) | Antibodies Incorporated/NeuroMab | 75–253 (RRID:AB_10675136) | Mouse | 1:1000 |
| LRRK2 pSer935 | MRC PPU Reagents and Services, University of Dundee | UDD2 10(12) (RRID:AB_2921228) | Rabbit | 1 µg/ml |
| LRRK2 pSer1292 | Abcam Inc. | ab203181 (RRID:AB_2921223) | Rabbit | 1:2000 |
| Rab10 pThr73 | Abcam Inc. | ab230261 (RRID:AB_2811274) | Rabbit | 1:1000 |
| Rab10 | Nanotools | 0680–100/Rab10-605B11 (RRID:AB_2921226) | Mouse | 1:1000 |
| GAPDH (6C5) | Santa-Cruz | sc-32233 (RRID:AB_627679) | Mouse | 1:5000 |
| Beta-actin | Cell Signaling Technology | #4967s (RRID:AB_330288) | Rabbit | 1:1000 |
| Alpha-Tubulin (DM1A) | Cell Signalling Technologies | #3873s (RRID:AB_1904178) | Mouse | 1:1000 |

**Table 3.**
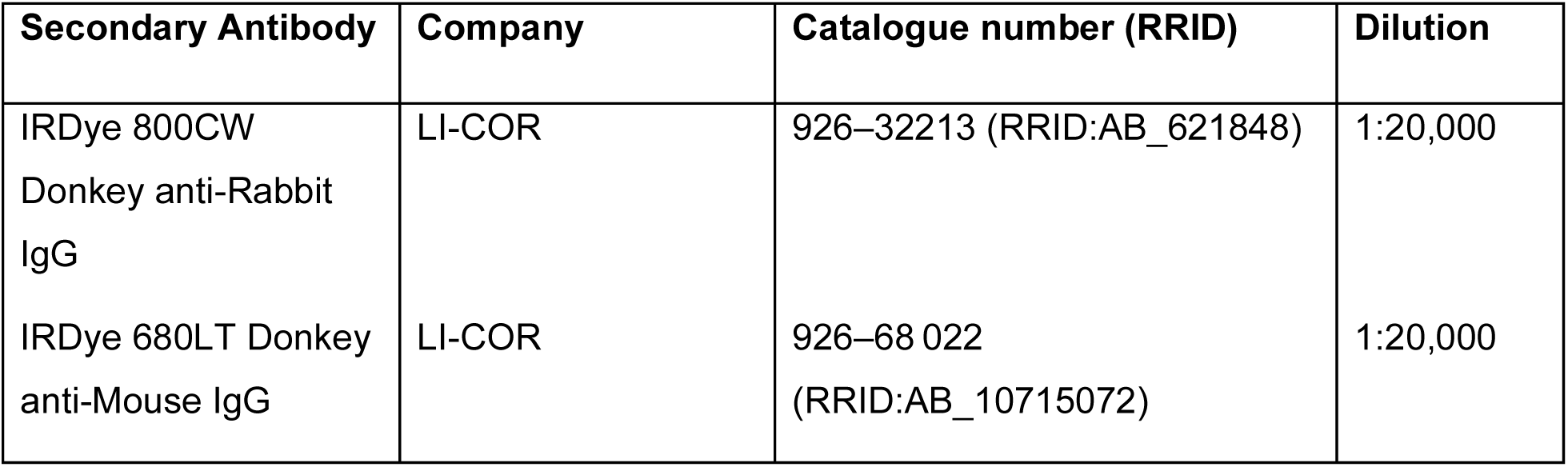
List of Secondary Antibodies used in this study.

#### Western Immunoblotting Data Analysis

Immunoblotting data (acquired using a LI-COR CLx Western Blot imaging system) were quantified using Image Studio Lite (Version 5.2.5, RRID:SCR_013715). Tubulin, β-actin or GAPDH were used as loading controls, as indicated in the respective figures. Phospho-Thr73-Rab10 and total Rab10, as well as phospho-Ser935-LRRK2 and total LRRK2, were detected by multiplex immunoblotting, with phosphorylation signals quantified relative to their corresponding total protein signals.

Quantified data were plotted with GraphPad Prism 10.5.0 (RRID:SCR_002798). For cell-based functional assays, data from three independent biological replicates were combined with values normalised to wild-type LRRK2 within each replicate.

For functional classification, variants producing a mean increase in Rab10 Thr73 phosphorylation of at least 1.5-fold relative to wild-type LRRK2 across a minimum of three independent biological replicates were considered to show increased pathway activity, as previously described (5, 15). This threshold was based on the magnitude of activation previously observed for the established pathogenic G2019S variant in the same assay system. Functional classification was independent of statistical significance, which was assessed using one-way ANOVA followed by Dunnett’s multiple-comparisons test.

#### Targeted PRM analysis for phospho-Rab12 antibody validation

Mouse embryonic fibroblasts (MEFs) carrying the VPS35[D620N] mutation, with or without RAB12 knockout, were cultured and treated with MLi-2 or DMSO as indicated.

For antibody validation, 100 µg of protein lysate from VPS35[D620N] MEFs treated with DMSO or MLi-2, and VPS35[D620N]/RAB12-knockout MEFs treated with DMSO (n = 4 per condition), was subjected to S-Trap-assisted digestion as previously described (https://www.protocols.io/view/sample-preparation-protocol-for-targeted-mass-spec-261ge4xoyv47/v1). The resulting peptides were vacuum-dried and resuspended in 800 µL of immunoaffinity purification (IAP) buffer supplemented with 100 fmol of stable isotope-labelled phospho-Rab12 peptide (mouse Ser105, corresponding to human Ser106). (10) Samples were incubated for 2 h with 0.5 µg of anti-phospho-Rab12 antibody (Abcam, ab325632; clone MJF-D29764-507) conjugated to protein A/G beads.

Beads were washed twice with 800 µL IAP buffer, twice with ice-cold PBS and once with ice-cold Milli-Q water. Phosphopeptides were eluted twice with 50 µL of 0.1% (v/v) trifluoroacetic acid (TFA), with each elution performed for 10 min at 1,200 rpm on a thermomixer. Eluates were recovered by centrifugation at 1,500 × g for 2 min, pooled and desalted using Evotips. Peptides were eluted from the Evotips in 20 µL of 50% (v/v) acetonitrile in 0.1% (v/v) formic acid, vacuum-dried and stored at −80 °C until analysis.

Peptides were resuspended in 6 µL of LC buffer containing 0.03% DDM, 3% (v/v) acetonitrile and 0.1% (v/v) formic acid. Targeted parallel reaction monitoring (PRM) was performed using an Orbitrap Astral mass spectrometer coupled to a Vanquish Neo LC system (Thermo Fisher Scientific). Peptides were separated on a 15 cm ES906 column using the 180 SPD method (approximately 7 min per run). MS1 spectra were acquired over an m/z range of 375–1,000 at a resolution of 120,000. Light and heavy phospho-Rab12 precursor ions (m/z 651.28 and 656.28, respectively; charge state 2+) were isolated using a quadrupole isolation width of 1.2 Da and fragmented by higher-energy collisional dissociation (HCD) at 27%. Fragment ions were detected in the Astral mass analyser over an m/z range of 100–1,400, with an AGC target of 100% and maximum injection times of 200 ms and 50 ms for light and heavy peptides, respectively. Raw data were processed using Skyline (version 25.1.1.206) and deposited with the ProteomeXchange Consortium via the PRIDE partner repository under dataset identifier PXD085777, and phospho-Rab12 abundance was quantified from light-to-heavy peptide peak-area ratios. Statistical comparisons between the three conditions were performed using one-way ANOVA in GraphPad Prism, with Benjamini–Hochberg correction for multiple testing at a false discovery rate of 5%.

#### Study participants and IRB approval

The study was approved by the Comité de Ética de la Investigación con Medicamentos (CEIm) at Hospital Universitario Fundación Alcorcón, Madrid, Spain. All participants provided written informed consent, and the study was conducted in accordance with the Declaration of Helsinki.

Participants were members of three generations of the Spanish family previously described in association with the LRRK2 p.[Leu119Pro;Leu488Pro] allele. (34)The study included 14 family members with and without Parkinson’s disease and while some participants were aware of their *LRRK2* genotype at recruitment, others had elected not to receive their genetic results. To minimise the risk of inadvertent genotype disclosure, participant numbers were randomly reassigned for figure presentation and do not correspond to the original study identifiers.

#### Genetic analysis

Family members underwent genetic testing to determine the *LRRK2* haplotype, including the p.L119P and p.L488P variants, and to assess the deletion of exons 4–11 of *GBA1*. The *LRRK2* variants were analysed using conventional PCR and Sanger sequencing. The primers used for p.Leu119Pro were forward 5′-CAC ATG AGC TTC ACT ACA G-3′ and reverse 5′-ATC AAG CTA CCC TAA TCC TG-3′; those used for p.Leu488Pro were forward 5′-GCC CTC CTG TAC TTA TTT CAA-3′ and reverse 5′-TGT TAT CTC TTC CCA CAA TCA-3′.

The *GBA1* gene was analysed using long-range PCR with high-fidelity DNA polymerase (Thermo Fisher Scientific), as previously described. (34) The primers were forward 5′-CGA CTT TAC AAA CCT CCC TG-3′ and reverse 5′-CCA GAT CCT ATC TGT GCT GG-3′. Amplicons were visualised on a 1% agarose gel.

To protect participant confidentiality and respect individual preferences regarding the disclosure of genetic results, GBA1 carrier status is not reported at the individual level.

#### Neutrophil and monocyte isolation

For analysis of LRRK2 pathway activity, 20 mL of fresh peripheral blood was collected by venepuncture from participating family members. Neutrophils and monocytes were isolated by immunomagnetic negative selection within 1 h of blood collection, as previously described (DOI: 10.1042/BCJ20170803; DOI: 10.1042/BCJ20180248). Cells were lysed in 5% SDS, 50 mM TEAB supplemented with protease and phosphatase inhibitors, 0.5 mM DIFP and 1 µg/µL microcystin-LR. Lysates were snap frozen and stored at −80 °C until further processing.

Detailed protocols from blood collection, cell isolation and lysis are available via protocols.io: https://dx.doi.org/10.17504/protocols.io.qpvonx8o9l4o/v1 and https://dx.doi.org/10.17504/protocols.io.jyl8nr646l2w/v1.

Lysates were shipped to Dundee for further processing, then thawed and sonicated, followed by centrifugation at 21,000 × g for 15 min at 4 °C. The clarified supernatants were collected, and protein concentrations were determined by BCA assay. Lysates were subsequently analysed by immunoblotting as described above or targeted parallel reaction monitoring (PRM) mass spectrometry, as described below.

#### Parallel reaction monitoring mass spectrometry for targeted LRRK2 pRabs analysis

For targeted proteomic analysis, 100 µg of total protein per sample was processed using an automated SP3-based workflow on an Agilent Bravo liquid-handling platform. Proteins were reduced and alkylated, captured onto SP3 beads, washed and digested with trypsin. Stable isotope-labelled heavy peptide standards corresponding to the targeted analytes were added to each sample. Phosphorylated Rab peptides and LRRK2 peptides were enriched by immunoprecipitation using a multiplexed antibody panel comprising a pan-pThr-Rab antibody recognising phosphorylated Rab1, Rab3, Rab8, Rab10, Rab35 and Rab43, a phospho-Rab12 (Ser106)-specific antibody, and antibodies against total LRRK2, pSer910-LRRK2 and pSer935-LRRK2. For total Rab analysis, a 5% aliquot of the tryptic digest was analysed separately without immunoenrichment.

Calibration curves were generated using synthetic light and stable isotope-labelled heavy peptides in an *E. coli* digest matrix. Each calibration sample contained 100 µg of *E. coli* digest and 100 fmol of the corresponding heavy peptide standards, together with a 12-point √10 serial dilution of light peptide standards.

Samples were loaded onto Evotips and analysed by parallel reaction monitoring (PRM) using an Evosep One LC system coupled to a Thermo Scientific Orbitrap Exploris 480 mass spectrometer. Total Rab peptides were analysed in a separate PRM run. PRM data were processed using Skyline [version 25.1.1.206]. Peptide abundance was determined from light-to-heavy peak-area ratios, with calibration curves used for absolute quantification. Phosphorylation levels were normalised to the corresponding total protein abundance, where applicable, while total LRRK2 was analysed based on protein abundance. Differences between LRRK2 variant carriers and non-carriers were assessed using Welch’s unpaired two-sided t-test on log10-transformed data, with Benjamini-Hochberg correction for multiple comparisons (FDR < 0.05).Statistical significance was defined as q < 0.05. Statistical analyses were performed using GraphPad Prism (Version 11.1.0). A detailed protocol is available on <u>protocols.io</u>: https://dx.doi.org/10.17504/protocols.io.5jyl839y8v2w/v1. The mass spectrometry proteomics data have been deposited with the ProteomeXchange Consortium via the PRIDE partner repository under dataset identifier PXD085751.

## Data Availability

Data used in the preparation of this article were obtained from the Global Parkinsons Genetics Program (GP2; https://gp2.org). Specifically, we used Tier 2 data from GP2 release 10 (https://doi.org/10.5281/zenodo.15748014). GP2 data can be requested through AMP PD (https://amp-pd.org). All code generated for this article, and the identifiers for all software programs and packages used, are available on GitHub (https://github.com/GP2code/GP2-global-genetic-variant-landscape) and archived on Zenodo (https://doi.org/10.5281/zenodo.15699539).
The mass spectrometry proteomics data have been deposited with the ProteomeXchange Consortium via the PRIDE partner repository under dataset identifiers PXD085751 (human neutrophils) and PXD085777 (mouse embryonic fibroblasts).
The experimental source data supporting the figures, including quantitative immunoblotting and associated analyses, are available via Zenodo (https://doi.org/10.5281/zenodo.23360970). Detailed experimental protocols are available on protocols.io and are cited in the corresponding Materials and Methods sections.
Individual-level clinical and genetic information from the participating family is not publicly available to protect participant confidentiality.

https://10.5281/zenodo.23360970

Supplementary Information is provided in the accompanying supplementary file.

## ACKNOWLEDGMENTS

The authors gratefully acknowledge the members of the family in Spain who participated in this study and generously contributed their time and samples. We acknowledge the excellent technical and laboratory support provided by the MRC Protein Phosphorylation and Ubiquitylation Unit (PPU), including laboratory management, the Cloning team, DNA Sequencing Service, tissue culture team, Mass Spectrometry Facility, and MRC PPU Reagents and Services teams. In particular, we thank Dr Fred Lamoliatte for his valuable contributions to the development and optimisation of the targeted parallel reaction monitoring (PRM) mass spectrometry assay and associated analytical protocol used in this study. The authors gratefully acknowledge Rob and Susan Whiteford for their outstanding fundraising efforts in support of this research programme, along with the many individuals who also participated. Their enthusiasm, generosity, and commitment have contributed significantly to the advancement of this work. This work was also supported by the UK Medical Research Council (MC_UU_00018/1) (D.R.A.) and The Michael J. Fox Foundation for Parkinson’s Research (MJFF) LRRK2 Investigative Therapeutic Exchange initiative (LITE), and by the joint efforts of the Aligning Science Across Parkinson’s (ASAP) initiative. MJFF administers the grant ASAP-000463 on behalf of ASAP and itself (to D.R.A). This study was also funded by Instituto de Salud Carlos III, project PI22/00680 (J.H.) and also by European Regional Development Fund “A way to make Europe” (J.H.).

This project was supported by the Global Parkinson’s Genetics Program (GP2; https://gp2.org), a research program of Aligning Science Across Parkinson’s (ASAP) (https://ror.org/03zj4c476).

ASAP is managed by the Coalition for Aligning Science (CAS) (https://ror.org/00rf4ez40). The Michael J. Fox Foundation for Parkinson’s Research (MJFF) (https://ror.org/03arq3225) is the funding and strategic partner. For a complete list of GP2 members see https://doi.org/10.5281/zenodo.7904831. For the purpose of open access, the author has applied a CC BY public copyright license to all Author Accepted Manuscripts arising from this submission.

## Data and Code Availability

Data used in the preparation of this article were obtained from the Global Parkinson’s Genetics Program (GP2; https://gp2.org). Specifically, we used Tier 2 data from GP2 release 10 (https://doi.org/10.5281/zenodo.15748014). GP2 data can be requested through AMP PD (https://amp-pd.org). All code generated for this article, and the identifiers for all software programs and packages used, are available on GitHub (https://github.com/GP2code/GP2-global-genetic-variant-landscape) and were given a persistent identifier via Zenodo (https://doi.org/10.5281/zenodo.15699539).

The mass spectrometry proteomics data have been deposited with the ProteomeXchange Consortium via the PRIDE partner repository under dataset identifier PXD085751 for the human neutrophil data and PXD085777 for the mouse embryonic fibroblast data.

The experimental source data supporting the figures, including quantitative immunoblotting, are available via Zenodo (10.5281/zenodo.23360970).

Detailed experimental protocols are available on protocols.io and are cited in the corresponding Materials and Methods sections.

Individual-level clinical and genetic information from the participating family is not publicly available to protect participant confidentiality.

## AUTHOR CONTRIBUTIONS

L.V.D., J.H., E.S. and D.R.A. conceived, designed and supervised the study. N.R. performed transient transfection and immunoblotting experiments, assisted by T.G. and A.G.A. (Figs. 1 and 4; Supplementary Fig. 2). S.G. performed PRM analysis, assisted by R.E.F. (Fig. 3). R.S. and E.S. isolated neutrophils and monocytes from patient blood, and T.M.G. performed immunoblotting analysis of neutrophil and monocyte extracts (Fig. 2). P.L. performed structural analysis (Fig. 1). L.M.L. and A.Z. provided information on other LRRK2 variants located near L488P. D.R.A. led the MJFF-funded phospho-Rab12 antibody development programme, with antibody screening by P.L., immunoprecipitation validation by R.N., and integration into multiplexed PRM by S.G. L.V.D. coordinated patient recruitment, and J.H. performed patient genetic analysis. All authors contributed to writing and review of the manuscript.

## Supplementary material

**Figure S1.**
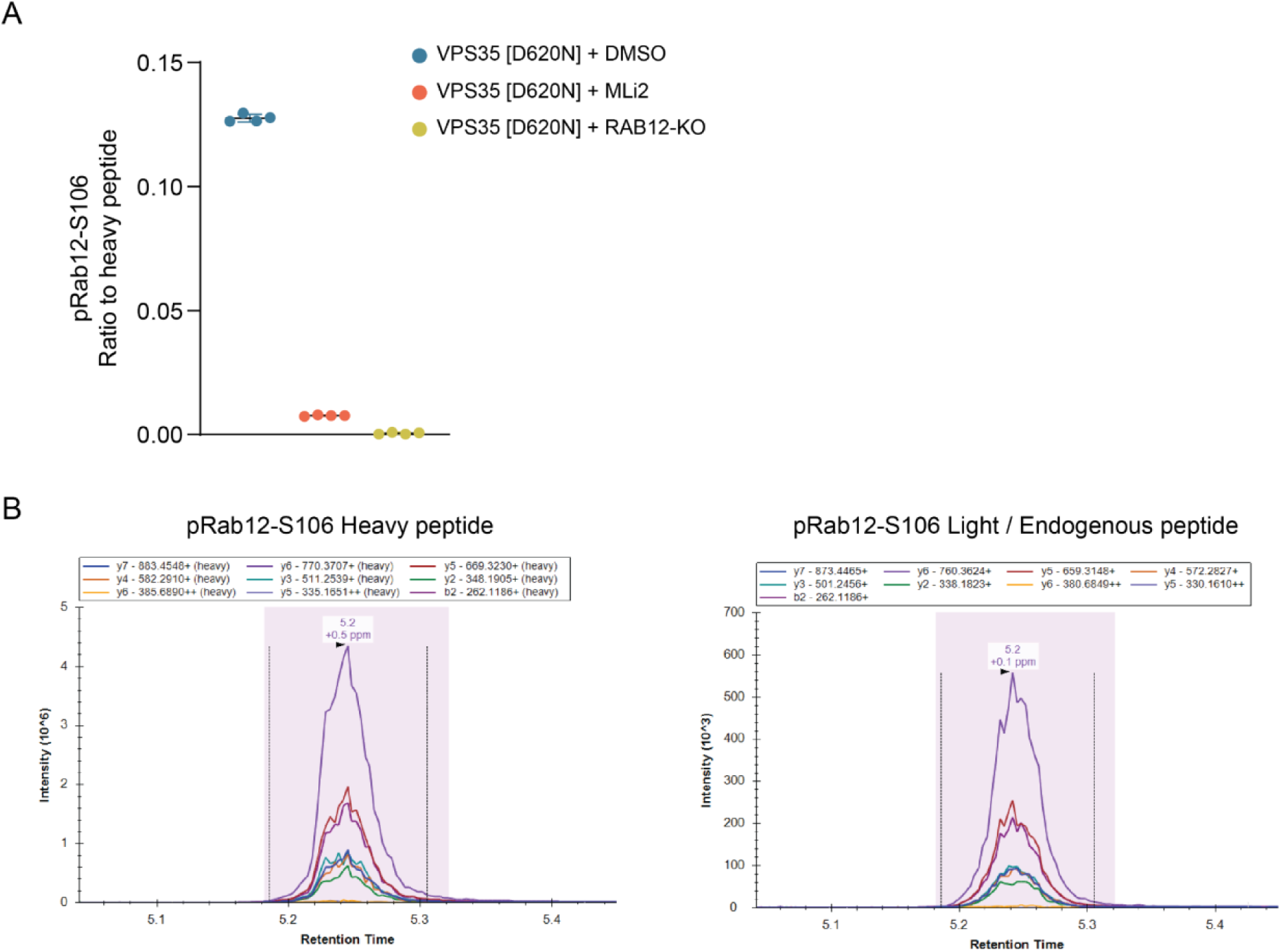
Validation of a phospho-Rab12 (Ser106) immunoprecipitating antibody. **(A)** Quantification of pRab12-S106 in VPS35[D620N] ± MLi-2 and VPS35[D620N]+Rab12-knockout mouse embryonic fibroblasts. Tryptic peptide digests were immunoprecipitated using a pRab12-S106 antibody (Abcam AB325632; clone MJF-D29764-507) and spiked with 100 fmol of heavy pRab12-S106 peptide and relative abundance presented as a ratio to heavy peptide between the conditions (n = 4 independent experiments; Significance: One-way ANOVA with Benjamini– Hochberg corrected 5% FDR). **(B)** Peak areas of heavy pRab12-S106 and endogenous or light pRab12-S106 peptide transition ions for the representative experiment shown in **(A)**.

**Figure S2.**
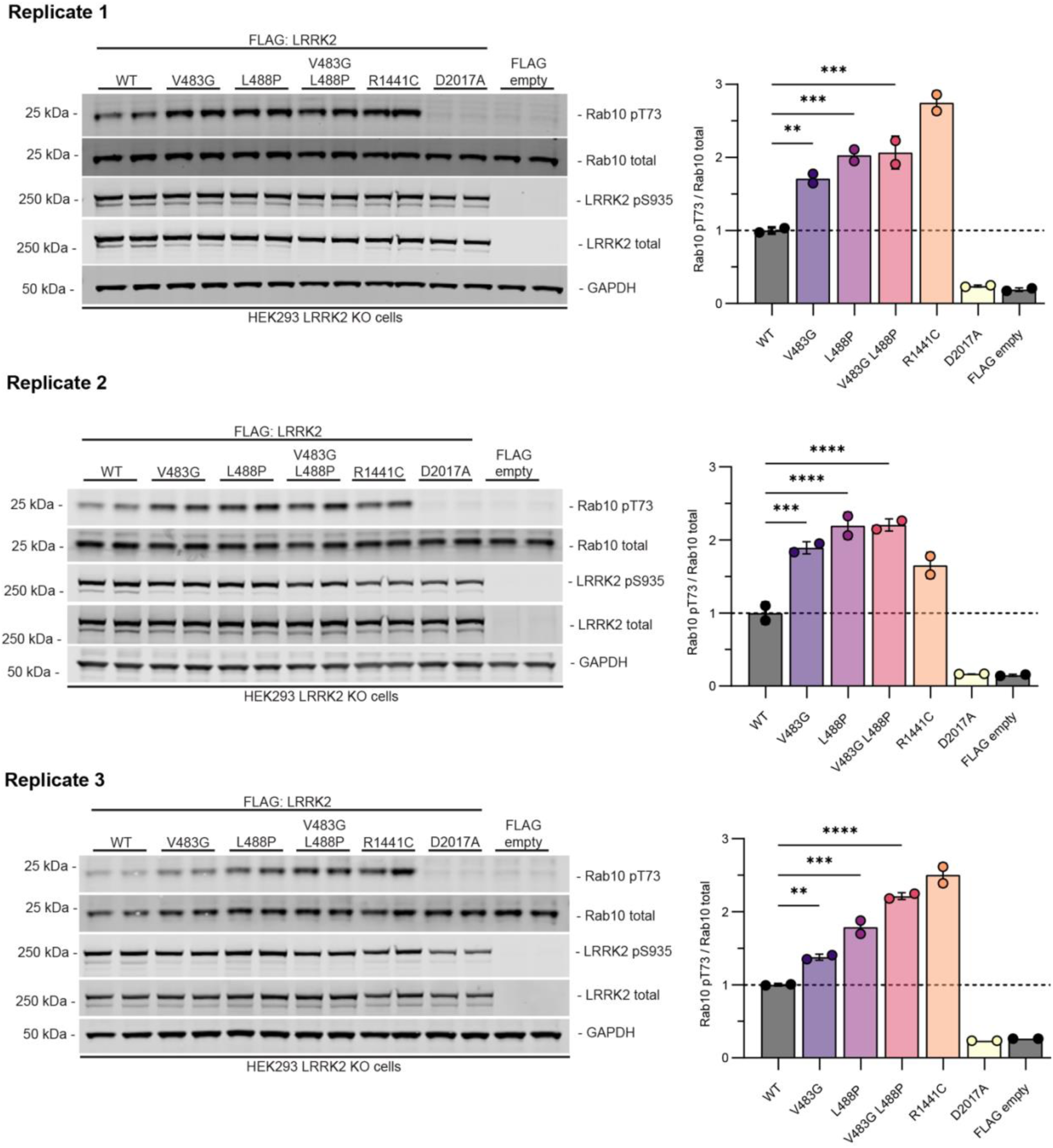
Combining V483G and L488P does not further enhance LRRK2 pathwayt activation. **(A)** Functional analysis of the indicated variants, including the V483G/L488P double variant, using transient overexpression of FLAG-tagged LRRK2 constructs in HEK293 LRRK2-knockout cells. Controls included wild-type (WT) LRRK2, the PD-associated R1441C variant (positive control), the kinase-dead D2017A variant (negative control), and an empty FLAG vector. Cells were lysed 24 h after transfection, and 30 µg of total protein per sample was analysed by quantitative immunoblotting with the indicated antibodies. The immunoblot analysis in panel A is representative of three independent biological replicates. **(B)** Quantification of Rab10 Thr73 phosphorylation from three independent biological replicates, expressed as the ratio of phospho-Thr73 Rab10 to total Rab10. Values within each replicate were normalised to the mean value for WT LRRK2 in that replicate. Data are presented as mean ± SD. Statistical analysis was performed using one-way analysis of variance (ANOVA tatistical analysis was performed using ordinary one-way ANOVA followed by Dunnett’s multiple-comparisons test versus WT. Asterisks indicate Dunnett-adjusted *P* values: P < 0.05 (*), P < 0.01 (**), P < 0.001 (***) and P < 0.0001 (****); ns, not significant. The dashed horizontal line indicates the normalised WT level (1.0). Source data are available on Zenodo (DOI: 10.5281/zenodo.23360970).

**Supplementary Table 1.** Statistical comparison of LRRK2 pathway measurements between L119P/L488P carriers and non-carriers. Individual LRRK2 pathway measurements were compared between five non-carriers and nine L119P/L488P carriers using two-sided unpaired Welch’s t-tests on log10-transformed data. P values were adjusted across ten measurements using the Benjamini–Hochberg false discovery rate (FDR) procedure at 5%.

| Measurement | Unadjusted P value | FDR-adjusted q value | Significant (FDR 5%) |
|---|---|---|---|
| pRab1 | 0.2258 | 0.2823 | No |
| pRab3 | 0.0041 | 0.0069 | Yes |
| pRab8 | <0.0001 | 0.0002 | Yes |
| pRab10 | 0.0003 | 0.0007 | Yes |
| pRab12 | 0.0003 | 0.0007 | Yes |
| pRab35 | 0.0004 | 0.0008 | Yes |
| pRab43 | 0.0003 | 0.0007 | Yes |
| pSer910-LRRK2 | 0.3663 | 0.4070 | No |
| pSer935-LRRK2 | 0.2151 | 0.2823 | No |
| Total LRRK2 | 0.4702 | 0.4702 | No |

Statistical significance was defined as q < 0.05. All six significantly increased phospho-Rab measurements remained significant after FDR correction. Values are reproduced as displayed in GraphPad Prism (four decimal places).

